# Humoral and cellular immune responses to CoronaVac assessed up to one year after vaccination

**DOI:** 10.1101/2022.03.16.22272513

**Authors:** Priscilla Ramos Costa, Carolina Argondizo Correia, Mariana Prado Marmorato, Juliana Zanatta de Carvalho Dias, Mateus Vailant Thomazella, Amanda Cabral da Silva, Ana Carolina Soares de Oliveira, Arianne Fagotti Gusmão, Lilian Ferrari, Angela Carvalho Freitas, Elizabeth González Patiño, Alba Grifoni, Daniela Weiskopf, Alessandro Sette, Rami Scharf, Esper Georges Kallas, Cássia Gisele Terrassani Silveira, the PROFISCOV study group

**Affiliations:** Medical Investigation Laboratory 60 (LIM-60), School of Medicine, University of São Paulo, São Paulo, SP, Brazil; Department of Infectious and Parasitic Diseases, Clinicas Hospital, School of Medicine, University of São Paulo, São Paulo, SP, Brazil; Butantan Institute, São Paulo, SP, Brazil; Center for Infectious Disease and Vaccine Research, La Jolla Institute for Immunology, La Jolla, CA 92037, USA; Department of Medicine, Division of Infectious Diseases and Global Public Health, University of California, San Diego, La Jolla, CA 92037, USA; PATH, Washington, DC 20001, USA

## Abstract

**Background:** The Sinovac SARS-CoV-2 inactivated vaccine (CoronaVac) has been demonstrated to be safe, well tolerated, and efficacious in preventing mild and severe Covid-19. Although different studies have demonstrated its short-term immunogenicity, long-term cellular and humoral response evaluations are still lacking.

**Methods:** Cellular and humoral responses were assessed after enrollment of volunteers in the PROFISCOV phase 3 double-blind, randomized, placebo-controlled clinical trial to evaluate CoronaVac. Assays were performed using flow cytometry to evaluate cellular immune response and an antigen binding electrochemiluminescence assay to detect antigen-specific antibodies to the virus.

**Results:** Fifty-three volunteers were selected for long term assessment of their SARS-CoV-2-specific immune responses. CD4^+^ T cell responses (including circulating follicular helper (cTfh, CD45RA^-^ CXCR5^+^) expressing CD40L, as well as non-cTfh cells expressing CXCR3) were observed early upon the first vaccine dose, increased after the second dose, remaining stable for 6-months. Memory CD4^+^ T cells were detected in almost all vaccinees, the majority being central memory T cells. IgG levels against Wuhan/WH04/2020 N, S and receptor binding domain (RBD) antigens and the variants of concern (VOCs, including B.1.1.7/Alpha, B.1.351/Beta and P.1/Gamma) S and RBD antigens peaked 14 days after the second vaccine shot, and were mostly stable for a 1-year period.

**Conclusions:** CoronaVac two-doses regimen is able to induce a potent and durable SARS-CoV-2 specific cellular response. The cellular reaction is part of a coordinated immune response that includes high levels of specific IgG levels against parental and SARS-CoV-2 VOC strains, still detected after one year.

**Funding:** Fundação Butantan, Instituto Butantan and São Paulo Research Foundation (FAPESP) (grants 2020/10127-1 and 2020/06409-1). This work has also been supported by NIH contract 75N93019C00065 (A.S, D.W). PATH facilitated reagent donations for this work with support by the Bill & Melinda Gates Foundation (INV-021239). Under the grant conditions of the foundation, a Creative Commons Attribution 4.0 generic License has already been assigned to the Author Accepted Manuscript version that might arise from this submission.

## Introduction

Protective and long-lasting immune responses induced by viral infections or vaccines are usually formed by a combination of humoral and cellular immunity. The presence of both SARS-CoV-2-specific responsive cells and circulating antibodies after infection or vaccination are the most likely candidates to serve as correlates of protection against symptomatic infections and reduction of disease severity. High frequency of circulating SARS-CoV-2-specific CD4^+^ and CD8^+^ T cells in patients who recovered from COVID-19, as well as the presence of memory T cells in the convalescent phase have been extensively described in literature and are suggested to play a key role in controlling SARS-CoV-2 initial infection, protection against virus re-infection and disease progression (Grifoni et al., 2020; Neidleman et al., 2020; Ni et al., 2020).

In 2020, Grifoni and colleagues described a functional validation of SARS-CoV-2 peptides megapools (MPs) in peripheral blood mononuclear cells (PBMCs) isolated from COVID-19 patients during the convalescence phase (Griffoni et al., 2020). They showed that these MPs were effective *in vitro* on identifying SARS-CoV-2-specific T cell responses, allowing the evaluation of antigen-specific CD8^+^ T cells, as well as multiple profiles of specific-CD4^+^ T cells induced by vaccination (Mateus et al, 2021).

CoronaVac, a COVID-19 vaccine administered to over 2 billion people (Dolgin, 2022), has a good safety profile and is effective against symptomatic SARS-CoV-2 infections and highly protective against moderate and severe COVID-19 (Bueno et al., 2021; Zhang et al., 2020). Brazilian phase 3 clinical trial of CoronaVac vaccine started on July 21, 2020 and enrolled 12,396 healthcare professionals that received the two-dose vaccine or placebo (Palacios et al., 2020). Among these volunteers, a 120-individuals cohort was selected for a prospective observational study to follow vaccine-induced cellular and humoral immune responses. We evaluated the humoral and cellular response in these volunteers and the data is presented here.

## Methods

### PROFISCOV phase 3 clinical trial

To assess the safety and efficacy of the CoronaVac vaccine in Brazil, a randomized, double-blind, placebo-controlled phase 3 multicenter clinical trial was performed in healthy healthcare professionals on the front line of COVID-19 patients’ treatment. The trial was approved by the Brazilian National Research Ethics Council (CONEP), CAAE 34634620.1.1001.0068, the Brazilian National Regulatory Agency (ANVISA), CE 47/2020, and was registered in the ClinicalTrials.gov platform (NCT04456595). The full protocol of the clinical trial has been published previously by Palacios et al. (2020). All participants provided written informed consent.

CoronaVac (Sinovac Life Sciences, Beijing, China) is an inactivated vaccine derived from the CN02 strain of SARS-CoV-2 grown in Vero cells. Production methods and its full composition and vaccination schedule has been published by Gao et al. (2020). The placebo group received aluminum hydroxide adjuvant with no virus. Vaccine and placebo were provided in a ready-to-use syringe and administered intramuscularly following the two-dose schedule of 0 and 14 days (Palacios et al., 2020).

From the 12,396 initial participants recruited in the Brazilian phase 3 clinical trial, 653 were included in the São Paulo site. From those, 120 individuals among vaccinees and placebos were randomly selected for a long-term follow-up and long-term assessment of their cellular and humoral responses.

### Study design and participants

After the breaking of the participants’ blinding code, these 120 individuals were filtered based on vaccination status, age and specimen availability to compose the cohort used to assess cellular and humoral response after immunization with CoronaVac. The final cohort was primarily composed of 29 vaccinees and 4 placebos ranging from 18 to 59 years, and 24 vaccinees and 4 placebos with ages over 60 years. Seven vaccinated volunteers were excluded from further analysis due to lack of follow-up or positive COVID-19 diagnosis right after the first vaccine dose. Placebos were only evaluated at day 0 and there were no infections reported in this group at this time point.

Blood samples were collected at eight time points starting at the day of the first dose until one year after vaccination (Fig. 1). The samples were processed in order to separate plasma and PBMCs, as described elsewhere (Paquin-Proulx et al., 2018). PBMCs and plasma were kept at liquid nitrogen and -80ºC, respectively, until further assays were performed.

**Figure 1.**
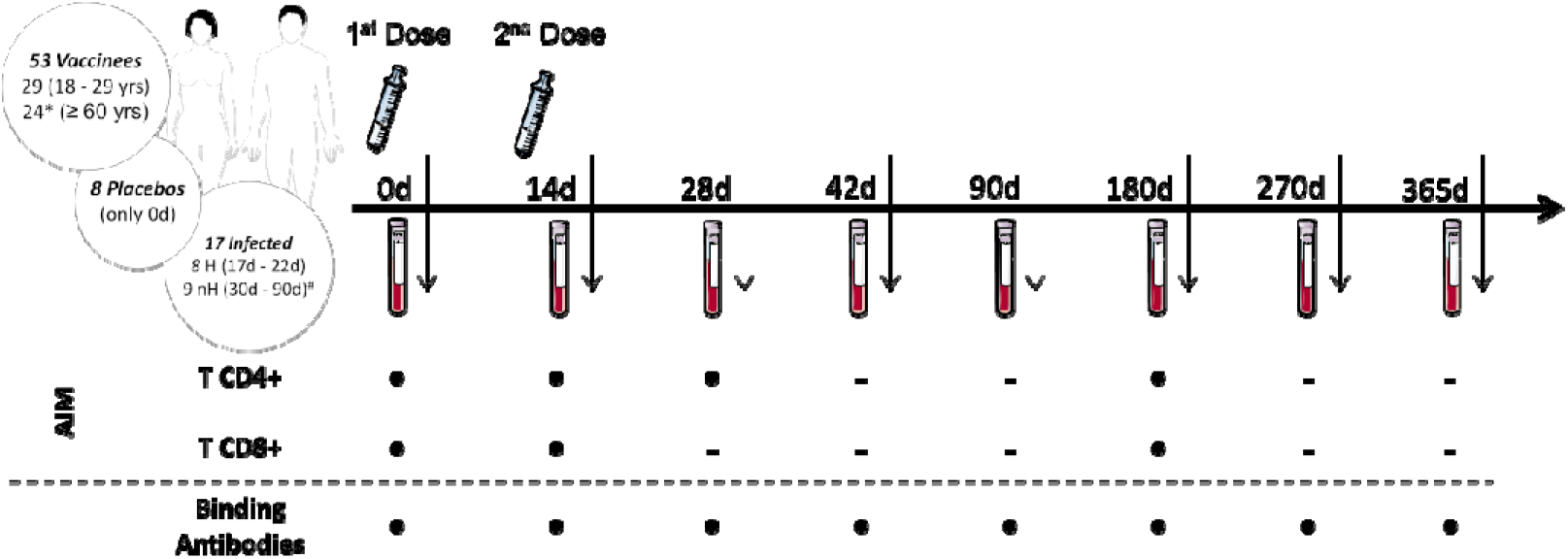
Experimental design of the volunteer cohort enrolled in the study. Each volunteer participating in the CoronaVac phase 3 clinical trial received either two doses of vaccine or placebo, with a 14-days median interval between doses. Blood samples from 0d and 14d were collected right before the first and second shot administration, from both vaccinees and placebos, respectively. As shown, 53 vaccinees were filtered for the analysis of antigenic-specific cellular response and binding antibody in blood samples collected at distinct time points as depicted in the timeline. Blood samples collected from 8 placebos at 0d and from 17 infected patients were added to balance the representation of healthy/unexposed and naturally infected individuals, respectively. AIM: Activation-Induced Markers assay. d: number of days after the first vaccine dose. H: Hospitalized COVID-19 infected patients. nH: non-Hospitalized COVID-19 infected patients. yrs: years of age. *One ≥60 vaccinee was excluded from the AIM assay due to lack of 0d PBMC sample. ^#^nH were only included in the binding antibody assay due to absence of PBMC samples.

In order to quantify the response magnitude achieved after vaccination, SARS-CoV-2 infected, age-paired individuals were selected as positive controls (eight hospitalized patients due to moderate COVID-19 symptoms and nine with mild symptoms that did not require hospitalization). Blood samples from these hospitalized patients are part of the SARS-CoV-2 neutralizing antibody investigation research project approved by CONEP, CAAE 34634620.1.1001.0068, and the non-hospitalized patients are infected placebos from the CoronaVac clinical trial. The demographics of the included individuals are described in Supplementary Table S1.

### Activation-induced markers (AIM) T cell assay

Evaluation of cellular parameters through activation-induced markers (AIM) assay included assessment of SARS-CoV-2-specific CD8^+^ and CD4^+^ T cell responses using virus-specific peptide pools developed by Grifoni and colleagues (2020). To surpass sensitivity limitations of cytokine-based assays, the AIM assay of CD4^+^ and CD8^+^ T cells was performed, which defines antigen specificity based on positive regulation of surface markers induced by T cell receptor (TCR) stimulation, instead of cytokine production. For this purpose, PBMCs from vaccinees and placebos were stimulated with MPs comprehending different portions of SARS-CoV-2, in order to quantify and determine the subsets of antigen-specific CD4^+^ and CD8^+^ T cells stimulated after the vaccine.

Briefly, PBMCs were thawed, counted and seeded at a density of 1,5×10^6^ cells/well. Cells were incubated with 0,5μg/mL anti-CD40 antibody (Miltenyi Biotec, NRW, Germany) and then stimulated in the presence of specific MPs (1μg/mL), 10μL/mL positive control phytohemagglutinin (PHA, Sigma-Aldrich, Darmstadt, Germany) or 0.1% negative control dimethyl sulfoxide (DMSO, Sigma-Aldrich, Darmstadt, Germany). MPs were developed and kindly donated by Dr. Alessandro Sette’s laboratory (Centre for Infectious Disease and Vaccine Research; La Jolla Institute for Immunology, USA) (Grifoni et al., 2020) and contain SARS-CoV-2-specific epitopes such as CD4-R (remaining non-Spike protein), CD4-S (Spike protein) and CD8-A and CD8-B (viral epitopes compatible with HLA-A and HLA-B most common genotypes, respectively). After incubation, cells were stained with a specific antibodies mix containing cell characterization and activation markers (Supplementary Table S2) (Moderbacher et al., 2020). After staining, cells were fixed with 1% paraformaldehyde (PFA, Sigma-Aldrich, Darmstadt, Germany) and resuspended in phosphate-buffered saline 1X (PBS, LGC Biotecnologia, SP, Brazil) for acquisition at the BD LSRFortessa™ X-20 Cell Analyzer (BD Biosciences, CA, USA). The full protocol is available at the Supplementary Appendix.

The quality control from samples acquired in the flow cytometer was performed by the analysis of the Fluorescence Minus One (FMO) and compensation adjustments through beads. Acquisition was performed with the BD FACSDiva™ Software v6.0 and FlowJo™ v10.8 was used for data analysis (both from BD Biosciences, CA, USA). T cell response values after viral peptide stimulus were obtained from the subtraction of the peptide-stimulated conditions from the DMSO condition. To generate data of total CD4^+^ and CD8^+^ T cells, separate responses for CD4-R and CD4-S and a sum of both were performed, as well for CD8-A and CD8-B.

### Binding antibody (bAb) assay

Plasma samples were tested for quantitative IgG bAbs against nine SARS-CoV-2 antigens: S, RBD and N from the Wuhan/WH04/2020 strain and S and RBD from the VOCs Alpha (B.1.1.7), Beta (B.1.351) and Gamma (P.1). An electrochemiluminescence multiplex serology assay (V-PLEX SARS-CoV-2 Panel 7 IgG kit, MesoScale Discovery (MSD), MD, USA) was used following manufacturer’s instructions. Plasma samples were heat inactivated at 56ºC for 45 minutes and used at a 1:1000 dilution.

Raw data was generated by Methodical Mind software (version 1.0.37; MSD) and analyzed with Discovery Workbench software (version 4.0; MSD). Antibody concentrations were calculated based on the 8-point calibration curve, specific for each one of the nine antigens, and reported as Arbitrary Units (AU/mL) adjusted by the dilution factor used in the assay. Detailed information concerning antigens, control samples and calibrators can be found in the kits inserts provided by MSD (cat. #K15437U).

### Statistical analysis

Graphs and statistical analysis were performed at GraphPad Prism version 9.0.0 for Windows (GraphPad Software, CA, USA). Non-parametric statistical tests Mann-Whitney and Kruskall-Wallis with Dunn’s correction for multiple comparisons were applied to compare groups, and differences were considered statistically significant for p values ≤0,05.

## Results

Vaccinee demographic analysis showed a median age of 36 years (IQR 31-42) and 55.2% female individuals in the 18-59 subgroup. The elder vaccinees (⍰60 years) had a median age of 67 years (IQR 62-70) and 66.7% were male individuals. COVID-19 hospitalized patients were predominantly male (87.5%) with a median age of 53 years (IQR 43-62) and a median age of 40 years (IQR 33-44) for non-hospitalized patients, who were 55.6% female. The placebo group had a median age of 54 years (IQR 36-64) and 62.5% were female individuals (Table S1).

### CoronaVac induces a 6-month lasting activated CD4^+^ response

Peptides designed to overlap the SARS-CoV-2 ORFeome enabled the assessment of SARS-CoV-2-specific CD4^+^ and CD8^+^ T cell immunity by a cytokine-independent *ex vivo* T cell assay in cryopreserved PBMC samples obtained prior and after vaccination. The total SARS-CoV-2-specific CD4^+^ T cell response (OX40^+^CD25^+^CD137^+^, Fig. 2A), presented as the sum values obtained for Spike and non-Spike CD4^+^ designed MPs, was detected 14 days after the first vaccine shot (p = 0.0001) (Fig. 2B). In five individuals (four from the 18-59 years group and one from the ⍰60 years group) SARS-CoV-2-specific CD4^+^ T cell response frequency reached more than 1% of total circulating CD4^+^ T cells with one vaccine dose, close to the frequency observed in some infected individuals (Fig. 2B). Higher frequencies were detected between 14 and 30 days after the second dose (28-42 days after first vaccine dose; p <0.0001) and this magnitude of response was maintained from 75 to 165 days after second dose (90 to 180 days after first dose; p <0.0001) (Fig. 2B). CD4^+^ T cells targeting the SARS-CoV-2 Spike ORF were the most representative cell subset of the total SARS-CoV-2-specific CD4^+^ T cell response at all time points evaluated after vaccination (p <0.0001) (Fig. 2C). CD4^+^ T cell responses to the non-spike SARS-CoV-2 ORFeome (MP-R) were found significant only at later time points (90 to 180 days after first dose; p <0.0032) (Fig. 2C). No significant SARS-CoV-2-specific CD8^+^ T cell responses were detected among vaccinees after the stimulation with SARS-CoV-2 MPs containing peptides targeting predicted CD8^+^ T cell epitopes of the most prominent HLA class I alleles (Fig. 2E).

**Figure 2.**
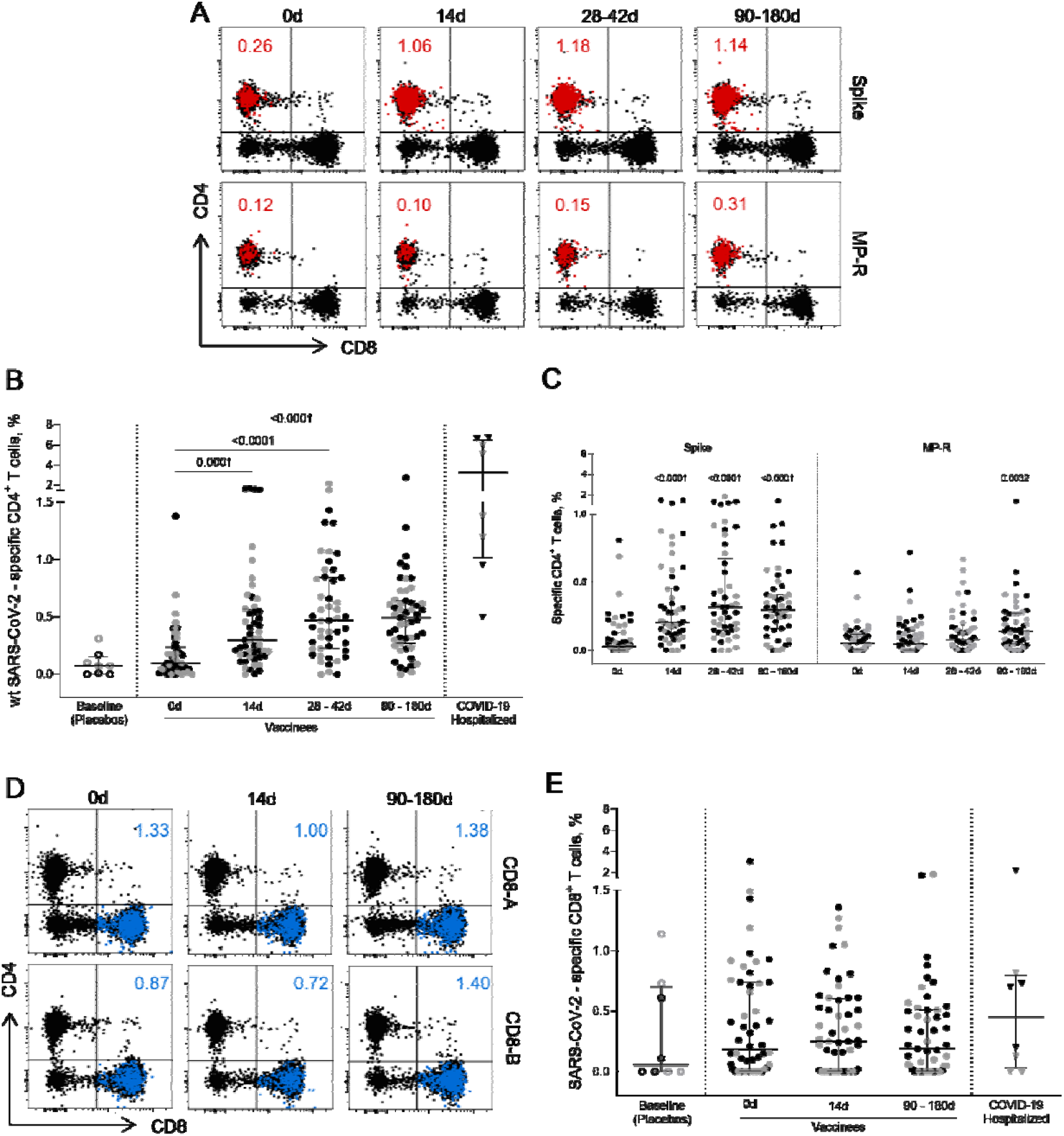
CD4^+^ and CD8^+^ circulating T cells activated after vaccination. (A) Representative FlowJo analysis of activated (OX40^+^CD25^+^CD137^+^) CD4^+^ T cells (in red) at four time points after the stimulation with SARS-CoV-2 Spike and non-spike (MP-R) peptides MPs. (B) Frequency of activated SARS-CoV-2-specific CD4^+^ T cells before and after first and second vaccine doses. (C) Frequency of activated SARS-CoV-2-specific Spike and non-Spike (MP-R) CD4^+^ T cells before and after the first and second vaccine doses. (D) Representative FlowJow analysis of activated (CD69^+^CD137^+^) CD8^+^ T cells (in blue) at three time points after stimulation with SARS-CoV-2 peptides MPs targeting most prominent HLA class I alleles (CD8-A and CD8-B) (E) Frequency of activated SARS-CoV-2-specific CD8^+^ T cells before and after the first and second vaccine doses. The percentage of activated cells was determined by using a Boolean gate on the total live CD4^+^ or CD8^+^ T cells. In the scatter dot plot graphs: symbol colours represent age-groups (black: 18-59, grey: ≥60); symbol shapes represent different volunteer sub-groups (unfilled circle: placebos (n =8), filled circle: vaccinees (n=53), filled triangles: SARS-CoV-2 infected hospitalized individuals (n=6), 19 days median of symptoms onset (IQR 17-22)). 0d: vaccinee baseline, day of first vaccine dose. 14d: two-weeks after the first dose, day of the second vaccine dose. Statistical comparisons using the Kruskall-Wallis test against baseline values were used.

### Tfh and nTfh cell subsets are elicited after first and second doses of CoronaVac

To assess SARS-CoV-2-specific CD4^+^ T cell response polarization, circulating T follicular helper (Tfh) and non-Tfh (nTfh) cells frequencies were measured based on the expression of the CXCR5 chemokine receptor. Both SARS-CoV-2-specific Tfh (CD45RA^-^CXCR5^+^) and nTfh (CD45RA^-^CXCR5^-^) cells frequencies had a significant increase after first vaccine dose (p =0.0076 and p <0.0001, respectively), showing a greater increment after the 14-day boost (p <0.0001; Figs. 3B and 3D). SARS-CoV-2-specific Tfh cell responses peaked 14 days after the second shot, subsequently dropping, although maintaining levels greater than those detected after the first vaccine dose (Fig. 3B).

**Figure 3.**
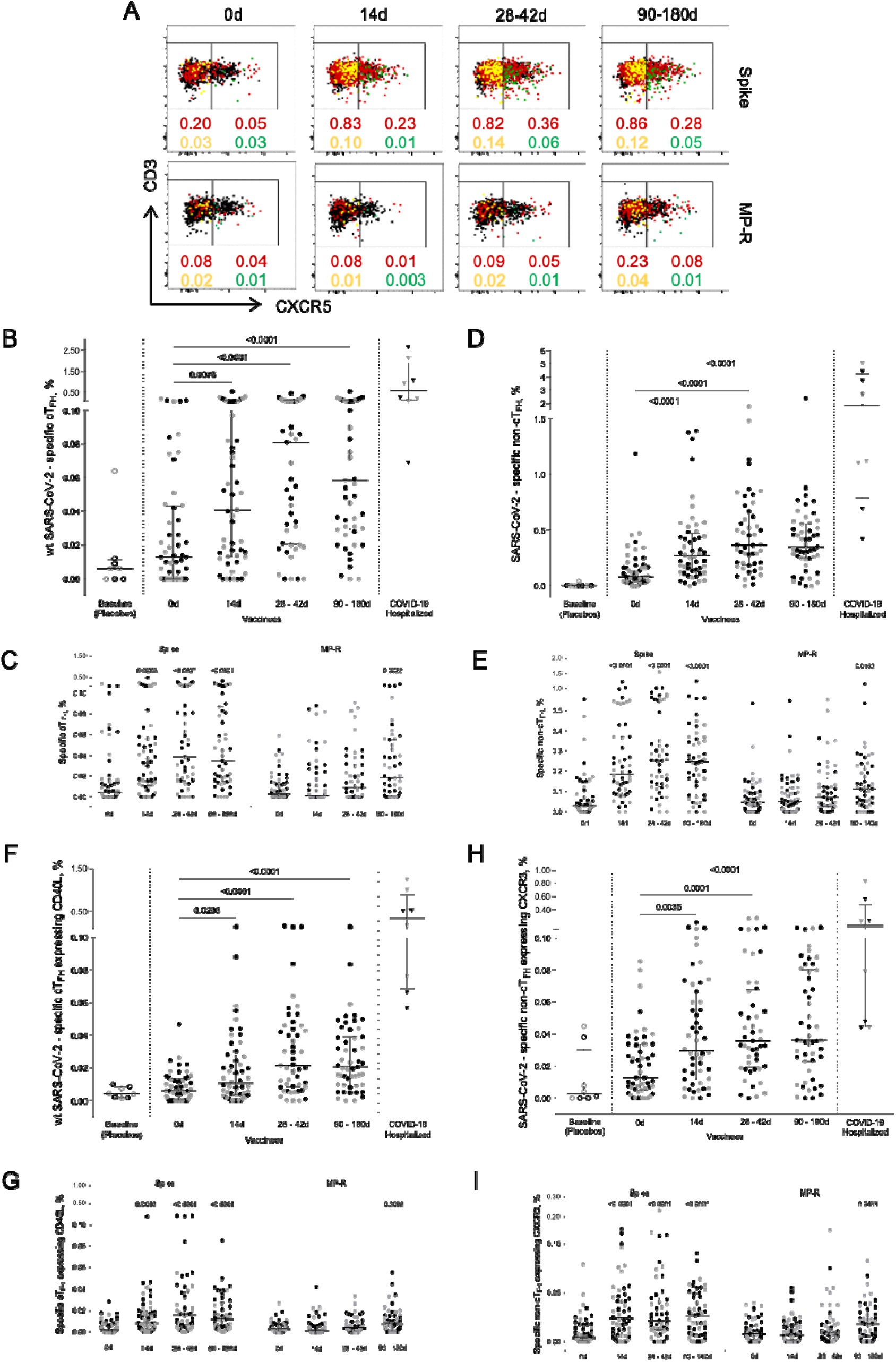
Specific SARS-CoV-2 Tfh and nTfh cells activation kinetics. (A) Representative FlowJow analysis strategy of activated (CD25^+^CD137^+^OX40^+^) Tfh (CD4^+^CD45^-^CXCR5^+^) and nTfh (CD4^+^CD45^-^CXCR5^-^) cells (in red) before and after the first and second vaccine doses. The yellow dots represent activated nTfh cells expressing CXCR3; the green dots represent activated Tfh cells expressing CD40L. (B) Frequency of activated SARS-CoV-2-specific Tfh cells. (C) Frequency of activated Tfh cells specific for SARS-CoV-2 Spike and MP-R peptides. (D) Frequency of activated SARS-CoV-2-specific nTfh. (E) Frequency of activated nTfh specific for SARS-CoV-2 Spike and MP-R peptides. (F) Frequency of activated SARS-CoV-2-specific Tfh expressing CD40L. (G) Frequency of activated Tfh expressing CD40L specific for SARS-CoV-2 Spike and MP-R peptides. (H) Frequency of activated SARS-CoV-2-specific nTfh expressing CXCR3. (I) Frequency of activated nTfh expressing CXCR3 specific for SARS-CoV-2 Spike and MP-R peptides. The percentage of cells co-expressing activation and other cell markers was determined by using a Boolean gate on the total live CD4^+^ T cells. In the scatter dot plot graphs: symbol colours represent age-groups (black: 18-59, grey: ≥60); symbol shapes represent different volunteer sub-groups (unfilled circle: placebos (n =8), filled circle: vaccinees (n =53), filled triangles: SARS-CoV-2 infected hospitalized individuals (n =6), 19 days median of symptoms onset (IQR 17-22)). 0d: vaccinee baseline, day of first vaccine dose. 14d: two-weeks after the first dose, day of the second vaccine dose. Statistical comparisons using the Kruskall-Wallis test against baseline values were used.

SARS-CoV-2-specific Tfh cells detected in post-vaccine time points were found to co-express CD40L, a T cell activation marker crucial to B cell maturation (Fig. 3F). Similar to what was previously observed for total CD4^+^ T cell response, Spike-specific Tfh circulating cells expressing CD40L were the most predominant cell subset detected after the two-dose vaccination and were significantly increased in all post-vaccine timepoints compared to baseline (14 days, p <0.003; 28-42 days and 90-180 days, p <0.0001, Fig. 3G). Significant CD40L^+^ Tfh cell MP-R-specific responses were detected after 90 days of the first vaccine shot (p =0.0099) (Fig. 3G). The bulk response against SARS-CoV-2 peptides for both Tfh and nTfh cells were directed by the Spike peptides MPs (Figs. 3C and 3E).

nTfh cells consist of heterogeneous cell populations that are often functionally stratified in humans by the expression of other chemokine receptors, including CXCR3 (Morita et al., 2011; Locci et al., 2013; Ueno, 2016). CXCR3^+^ nTfh are Th1-polarized cells and they were significantly increased after the first vaccine dose (p <0.0001) (Fig. 3H). Over 70% of the vaccinees had the frequency of CXCR3^+^ nTfh cells specific for SARS-CoV-2 above median values detected at baseline and the majority of this response was driven by Spike-peptides (Fig. 3I).

### Activated memory CD4^+^ cells are seen early on after first vaccine dose and are at least 3 months stable

Circulating SARS-CoV-2-specific memory CD4^+^ T cells (i.e. non-naïve CD4^+^CD45RA^+^CCR7^+^ T cells) were identified at a quite high rate early on after one vaccine dose (p <0.0001), and this frequency was increased and maintained over a six-month length period (p <0.0001) (Fig. 4B). The expansion of circulating SARS-CoV-2 memory CD4^+^ T cells (above the baseline median value) at 14 days after the first vaccine dose was found in 83% (43/52) of vaccinees. Between 28 and 42 days after the second dose the percentage of vaccinees with SARS-CoV-2 memory CD4^+^ T cells above the median basal level increased to 92% (45/49) and was maintained at this level in 88% of the individuals between 90 and 180 days (45/51). Memory T CD4^+^ cells activation was mainly triggered by Spike-derived peptide stimulation. The frequency of MP-R specific memory CD4^+^ cells significantly increased only at the latest time point evaluated (90 to 180 days after first shot, p =0.0030) (Fig. 4C).

**Figure 4.**
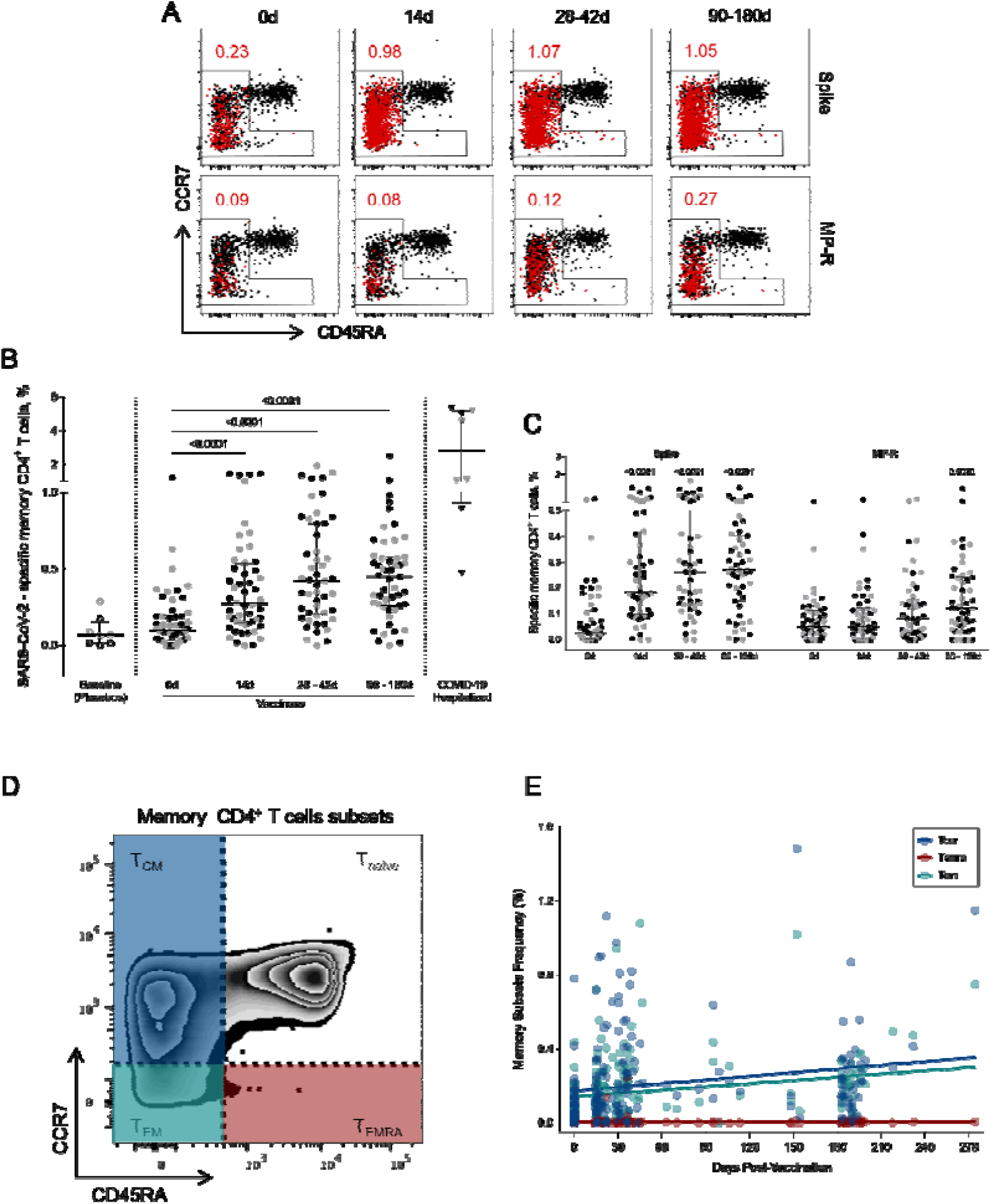
Activated memory CD4^+^ cTfh specific for SARS-CoV-2 peptides. (A) Representative FlowJow analysis of activated (CD25^+^CD137^+^OX40^+^) memory T CD4^+^ T cells (in red) at four time points. Memory CD4^+^ T cells here encompass all cell subsets based on surface expression of CD45RA and CCR7 except the naive subset (CD45RA^+^ CCR7^+^) (B) Frequency of activated memory CD4^+^ T cells specific for SARS-CoV-2 before and after the first and second vaccine doses. (C) Frequency of activated memory CD4^+^ T cells specific for SARS-CoV-2 Spike and MP-R peptides. (D) FlowJow analysis strategy example showing the subsets of memory CD4^+^ T cells: central memory (T_CM_) (CCR7^+^CD45RA^-^) in blue; effector memory (T_EM_) (CCR7^-^ CD45RA^-^) in green; and terminally differentiated effector memory cells (T_EMRA_) (CCR7^-^CD45RA^+^) in red. (E) Distribution of the activated memory CD4^+^ T cell subsets among total SARS-CoV-2-specific CD4^+^ T cells. The percentage of cells co-expressing activation and other memory cell markers was determined by using a Boolean gate on the total live CD4^+^ T cells. In the scatter dot plot graphs: symbol colors represent age-groups (black: 18-59, grey: ≥60) and symbol shapes represent different volunteer sub-groups (unfilled circle: placebos (n =8), filled circle: vaccinees (n =53), filled triangles: SARS-CoV-2 infected hospitalized individuals (n =6), 19 days median of symptoms onset (IQR 17-22)). 0d: vaccinee baseline, day of first vaccine dose. 14d: two-weeks after the first dose, day of the second vaccine dose. Statistical comparisons using the Kruskall-Wallis test against baseline values were used.

To further characterize the SARS-CoV-2-specific memory CD4^+^ T cells detected after vaccination, we assessed the diversity of memory cell subsets (Fig. 4D). Central memory (T_CM_) and effector memory (T_EM_) cells appear to be the most representative memory cell subsets due to progressive frequency increase after first and second vaccine doses while no change was observed in terminally differentiated effector memory cells (T_EMRA_) (Fig. 4E).

### CoronaVac induces high levels of specific IgG against wt SARS-CoV-2 and VOCs that are stable for at least 1-year period

As long-lasting SARS-CoV-2-specific Tfh cells were detected in the AIM assay, the next goal was to assess the levels of circulating antibodies against SARS-CoV-2 proteins in adult and elderly vaccinees, up to one year after the first vaccine dose. Analysis of both groups taken together showed a significant increase in the anti-Wuhan/WH04/2020-SARS-CoV-2 IgG titers 14 days after the first vaccine shot for Spike and RBD proteins but not for the N antigen. Two weeks after the complete vaccination scheme IgG levels against wt-SARS-CoV-2 N, Spike and RBD proteins were significantly higher compared to baseline levels (p <0.0001, Fig. 5A-C). Through one year follow-up SARS-CoV-2 N-specific IgG titers were relatively stable from 28 to 90 days after first vaccine shot, going gradually down in the next 6 months until one year after vaccination (Fig. 5A). Wuhan/WH04/2020-SARS-CoV-2 S- and RBD-specific IgG titers peaked from 28 to 42 days at similar levels found in non-hospitalized individuals (46 days median after symptoms onset, IQR 34-65) (Figs. 5B-C). Between 90 and 270 days S- and RBD-specific IgG levels showed a slight decay, reaching stability point from 270 to 365 days after first vaccine dose at significantly higher levels compared to baseline (p <0.0001) and placebo group (p <0.0001) (Fig. 5B-C).

**Figure 5.**
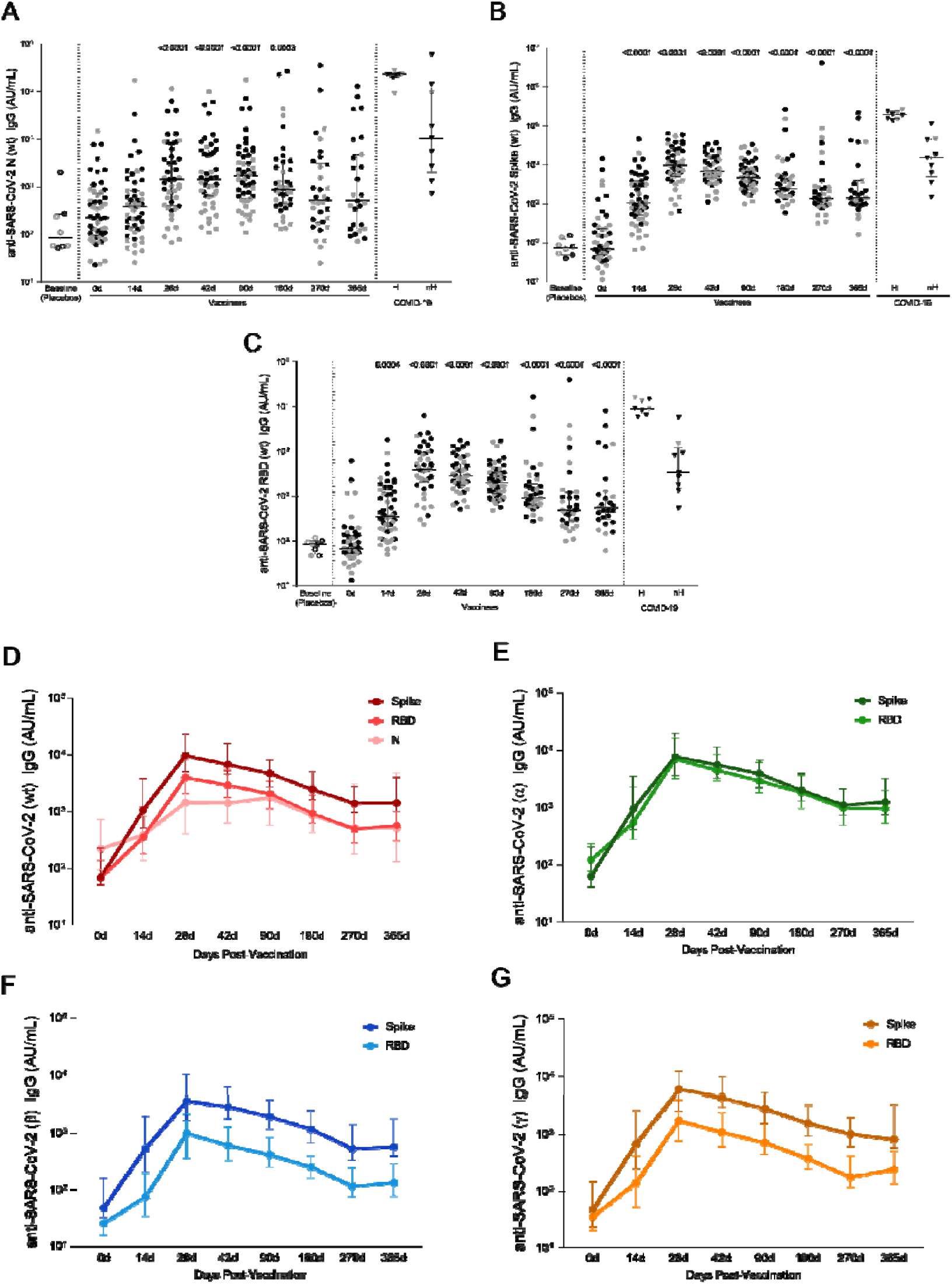
Specific wt-SARS-CoV-2 and VOCs strains IgG titers. (A) IgG levels measured against Nucleocapsid, (B) Spike and (C) RBD proteins from the Wuhan/WH04/2020-SARS-CoV-2 (wt). (D) IgG titers against S, RBD and N proteins from the Wuhan/WH04/2020-SARS-CoV-2 (wt). IgG levels against S and RBD proteins from (E) Alpha (B.1.117), (F) Beta (B.1.351) and (G) Gamma (P.1) strains. In the scatter dot plot graphs: symbol colors represent age-groups (black: 18-59, grey: ≥60) and symbol shapes represent different volunteer sub-groups (unfilled circle: placebos (n =8), filled circle: vaccinees (n =53), filled triangles: SARS-CoV-2 infected hospitalized (n =6) (19 days median of symptoms onset (IQR 17-22)) or non-hospitalized individuals (n =6) (46 days median of symptoms onset (IQR 34-65)). 0d: vaccinee baseline, day of first vaccine dose. 14d: two-weeks after the first dose, day of the second vaccine dose. In (D), (E), (F) and (G) values are expressed as the median and the 25-75% IQR. AU: arbitrary units. Statistical comparisons using the Kruskall-Wallis test against baseline values were used.

The same IgG response profile was found when analyzing antibodies against S and RBD proteins of Alpha, Beta and Gamma VOCs (Figs. 5E-G). IgG response peaked at 28 days after first vaccine dose, and even though it subsequently presented a slight decrease, antibody levels remained significantly higher after one year of vaccination compared to baseline (median value >10^2^ AU/mL, and p <0,0001 for all strains, Fig. S1, Table S3). It should be noted that the method used to calculate anti-Spike, anti-RBD and anti-N IgG titers against Wuhan/WH04/2020 and VOCs is based on an unique standard curve for each variant peptide target, and they cannot be directly compared. Nevertheless, the data shows that all titers have a similar expansion kinetics profile (Figs. 5D-G).

By assessing the Wuhan/WH04/2020-SARS-CoV-2 and VOCs IgG responses in each age group, a higher level of antibodies against S and RBD proteins were found among adult vaccinees when compared to the elderly individuals between 14 to 42 days after first vaccine shot. From 90 to 365 days after first dose, similar Wuhan/WH04/2020-SARS-CoV-2 and VOCs IgG levels were observed between both age groups (Fig. S1). No significant increase in IgG response against Wuhan/WH04/2020-SARS-CoV-2 N protein was detected among elderly vaccinees, whereas in vaccinated adults significant N-specific IgG levels were found between 28 and 180 days after first dose (Table S3).

## Discussion

COVID-19 vaccines that elicit protective immune responses are essential to prevent and mitigate the morbidity and mortality caused by SARS-CoV-2 infection. The aim of this study was to evaluate the immune response to CoronaVac by measuring the T cell activation kinetics after re-stimulation with SARS-CoV-2 peptides and the IgG-specific levels against the Wuhan/WH04/2020 strain and VOCs antigens. The results add understanding on how this vaccine builds and maintains the immune response.

Multiple lines of evidence support the critical roles of T cells in mounting immune responses to COVID-19. It is well-known that T cells can engage several antigen epitopes providing a broader protection against the virus (Zhao et al., 2016). SARS-CoV-2-specific peptides, which have been recognized by CD4^+^ and CD8^+^ T cells of exposed donors (Grifoni et al., 2020), allowed the detection of specific T cell responses even in individuals without detectable antibody responses, thereby providing evidence for T cell immunity upon vaccination. In line with previously published data showing an important role of SARS-CoV-2 CD4^+^ T cell responses in individuals recovering from COVID-19 infection (Grifoni et al., 2021) and after mRNA vaccination (Painter et al., 2021), a significant CD4^+^ T cell response triggered by Spike and non-Spike SARS-CoV-2 peptides was detected in peripheral blood cells of a subset of adults and elderly subjects vaccinated with CoronaVac. The data shown here indicates that CD4^+^ T cell may target different SARS-CoV-2 epitopes and suggest that the presence of additional SARS-CoV-2 antigens, such as M and N, in future vaccine formulations would enable it to better mimic the SARS-CoV-2-specific CD4^+^ T cell response seen in naturally infected patients.

A significant percentage of the circulating SARS-CoV-2-specific CD4^+^ T cells detected after two doses of CoronaVac exhibited a Tfh phenotype, similar to those observed following mRNA vaccination (Painter et al., 2021) and infection (Juno et al., 2020). Tfh cells are CD4^+^ lymphocytes specialized in regulating the adaptive immune response in germinal centers by enabling the selection of specific high affinity B cells and modulating affinity maturation in infection and vaccination (Crotty, 2014). Therefore, Tfh cells are crucial for establishing durable humoral immunity, by helping structuring antibodies generation. Mudd and colleagues (2021) demonstrated a significant correlation between the size of the germinal center B cell population in lymph nodes and the total Tfh cell population frequency following mRNA vaccination. Using a Spike immunodominant epitope, the authors showed an increase of specific Tfh in the blood of vaccinated individuals, peaking 28 days after the first dose. Interestingly, in our study we observed that not only circulating Tfh cells, but also the antibodies titers against Wuhan/WH04/2020-SARS-CoV-2 Spike and RBD antigens peaked between 28 and 42 days after CoronaVac first dose.

As circulating antibodies titres against Wuhan/WH04/2020 and VOCs strains reached its highest on day 28 after first dose, we showed that bAb (binding antibody) against Wuhan/WH04/2020 Spike and RBD proteins had at least 1 year durability, a profile also seen post-infection, but not with such longevity (Dan et al., 2021), and had also a similar median range as identified in our non-hospitalized cohort with 46 days post-symptoms onset. Although antibodies against the N protein showed a relevant decrease after 6 months, not seen in infected individuals (Dan et al., 2021), about half of the individuals remained above the median levels until 1 year after first vaccine dose. To our knowledge this is the first study to provide such a long-lasting bAb response for vaccinated individuals. A similar kinetics was observed for the bAb against all Spike and RBD VOCs (Alpha, Beta and Gamma) proteins levels when its decreasing stopped at 9 months and remained stable still above the basal IgG level throughout 1 year after first dose, different from the diminishing tendency seen in another study for the IgG levels against Beta, Gamma and Delta Spike targets in non-hospitalized individuals after 4 months of COVID-19 diagnose (Jodaylami et al., 2021).

Stratifying the age groups, we noticed that elderly (≥60 years) vaccinees delayed to reach significant bAb levels for wt and VOCs, kept at lower levels compared to those seen in adults (18-59 years) and dropped at earlier time points. Delayed and lower magnitude of antibody response in elderly individuals has been associated with the diminished frequency and response of naive circulating B and T cells in elderly individuals (Linton PJ, Dorshkind K., 2004).

T cells also have a direct role in clearing virus infections. In addition to help B cell and antibody responses, eliciting broad and long-lasting antiviral immunity requires the enrollment of CD4^+^ T cells and the generation of effective T cell memory (Le Bert et al., 2020) essential for protection against future infections. SARS-CoV-2 memory CD4^+^ T cells were detected in almost all subjects after receiving the two-dose regimen of CoronaVac, as well as in a cohort of mostly non-hospitalized patients with COVID-19 evaluated in another study (Dan et al., 2021). In contrast to the memory subset proportion (T_CM_ followed by T_EM_ subset along the 6 months) seen here after CoronaVac vaccination and also after mRNA vaccination (Painter et al., 2021), COVID-19 infected individuals showed an inverse proportion, being T_EM_ the most frequent for 6 months, being outpassed by the T_CM_ subset after this timeframe (Dan et al., 2021). These data suggest that vaccination with CoronaVac elicits an early sustained memory immunity compared to infection, since T_EM_ cells are associated with a more immediate defense.

In summary, the results shown herein provide evidence that CoronaVac is capable to induce a long-term antigen-specific CD4^+^ T cell response and develop high-titer antibody responses against Wuhan/WH04/2020 SARS-CoV-2 and VOCs strains in adults and elderly individuals and also highlight the importance of the two-dose regimen to establish a robust cellular and humoral response.

## Data Availability

All data produced in the present work are contained in the manuscript

## Acknowledgements

We would like to express our thanks to the study subjects for their participation and desires to forward our scientific work. We would also like to acknowledge all individuals from the PROFISCOV study group, specially Wellington Briques (MD, MBA) and Gecilmara Salviato Pileggi (MD, MSc, PhD), for the scientific and technical assistance. This work has been supported by Fundação Butantan, Instituto Butantan and São Paulo Research Foundation (FAPESP) (grants 2020/10127-1 and 2020/06409-1). This work was additionally supported by NIH contract 75N93019C00065 (A.S, D.W). PATH facilitated reagent donations for this work with support by the Bill & Melinda Gates Foundation (INV-021239).

