## Supplemental Files for "Humoral and cellular immune responses to CoronaVac assessed up to one year after vaccination"

### SUPPLEMENTARY APPENDIX


Activation-induced markers (AIM) T cell assay....................................................... 1

List of Reagents………………………………………………………………….. 2

Figure S1........................................................................................................ 3

Table S1. Demographic data of the study's participants.…………………..................... 7

Table S2. Antibodies and fluorochromes used in the activation induced marker (AIM) assay……......................................................................................................... 8

Table S3. Statistical differences on IgG levels against wild-type SARS-CoV-2 and SARS-CoV-2 variants of concern (VOCs) …………….......................................... 9

**Activation-induced markers (AIM) T cell assay - full protocol**

PBMCs were thawed at 37ºC and diluted in RPMI 1640 media (Thermo Scientific, MA, USA) supplemented with 5% human serum (Sigma-Aldrich, Darmstadt, Germany), 2 mM L-glutamine (Gibco, MA, USA), 1 mM penicillin/streptomycin (Gibco, MA, USA), 10 mM HEPES solution (Gibco, MA, USA), 1mM sodium pyruvate (Gibco, MA, USA) and 55 mM β-mercaptoethanol (Gibco, MA, USA) (hereafter referred as HR5). Cells were centrifuged at 1500 rpm for 10 min, resuspended in HR5 containing 50 U/mL benzonase (Merck, Darmstadt, Germany) and incubated at 37ºC and 5% $CO_2$ to remove DNA/RNA aggregates. After incubation, cells were counted, centrifuged at 1500 rpm for 10 min and seeded at a density of $1,5x10^6$ cells per well, in U-bottom 96-well plates.

Cells were incubated with 0,5 µg/mL anti-CD40 antibody (Miltenyi Biotec, NRW, Germany) for 15 min at 37ºC and 5% $CO_2$ and then stimulated for 24h at 37ºC and 5% CO2 in the presence of specific MPs (1 µg/mL), phytohemagglutinin (PHA, Sigma-Aldrich, Darmstadt, Germany) (10 µL/mL) as a positive control or dimethyl sulfoxide (DMSO, Sigma-Aldrich, Darmstadt, Germany) (0,1 %) as a negative control, all diluted in HR5. MPs were developed and kindly donated by Dr. Alessandro Sette's laboratory (Center for Infectious Disease and Vaccine Research; La Jolla Institute for Immunology, USA) (Grifoni et al., 2020) and comprehend SARS-CoV-2-specific epitopes specified as CD4-R (remaining non-Spike protein), CD4-S (Spike protein) and CD8-A and CD8-B (viral epitopes compatible with HLA-A and HLA-B, respectively).

After a 24-hours MPs incubation, plates were centrifuged at 1800 rpm for 2 min and the supernatant was collected in a new plate and kept at -80ºC freezer for further analysis. Cell pellets were resuspended, transferred to V-bottom 96-well plates and washed with MACS buffer (5 mg/mL bovine serum albumin (BSA, Sigma-Aldrich, Darmstadt, Germany) and 2 mM ethylenediamine tetraacetic acid (EDTA, Sigma-Aldrich, Darmstadt, Germany) diluted in phosphate-buffered saline 1X (PBS, LGC Biotecnologia, SP, Brazil)) twice, then stained with a specific antibodies mix containing characterization and activation markers (Supplementary Appendix) for 20 min, at 4ºC in the dark (Moderbacher et al., 2020). After staining, cells were washed twice at the same conditions, fixed with paraformaldehyde (PFA, Sigma-Aldrich, Darmstadt, Germany), diluted 1:10 in PBS 1X, for 10 min at room temperature in the dark, centrifuged at 2000 rpm for 5 min, resuspended in PBS 1X and acquired at the BD LSRFortessa™ X-20 Cell Analyzer (BD Biosciences, CA, USA).

**List of Reagents**

1. RPMI 1640 media (Thermo Scientific, MA, USA)
2. 5% human serum (Sigma-Aldrich, Darmstadt, Germany)
3. 2 mM L-glutamine (Gibco, MA, USA)
4. 1 mM penicillin/streptomycin (Gibco, MA, USA)
5. 10 mM HEPES solution (Gibco, MA, USA)
6. 1mM sodium pyruvate (Gibco, MA, USA)
7. 55 mM β-mercaptoethanol (Gibco, MA, USA)
8. 50 U/mL benzonase (Merck, Darmstadt, Germany)
9. Specific SARS-CoV-2 megapools (Center for Infectious Disease and Vaccine Research; La Jolla Institute for Immunology)
10. anti-CD40 antibody (Miltenyi Biotec, NRW, Germany)
11. Phytohemagglutinin (PHA, Sigma-Aldrich, Darmstadt, Germany)
12. Dimethyl sulfoxide) (DMSO, Sigma-Aldrich, Darmstadt, Germany)
13. U-bottom 96-well culture plates (TPP, Trasadingen, Switzerland)
14. MACS buffer:
    -5 mg/mL bovine serum albumin (BSA, Sigma-Aldrich, Darmstadt, Germany)
    -2 mM ethylenediamine tetraacetic acid (EDTA, Sigma-Aldrich, Darmstadt, Germany)
    - phosphate-buffered saline 1X (PBS, LGC Biotecnologia, SP, Brazil)
15. Paraformaldehyde (PFA, Sigma-Aldrich, Darmstadt, Germany), diluted 1:10 in PBS 1X
16. V-bottom 96-well plates (NUNC, Thermo Scientific, MA, USA)
17. V-PLEX SARS-CoV-2 Panel 7 IgG kit, catalogue number K15437U-4 (MesoScale Discovery, MD, USA)

A

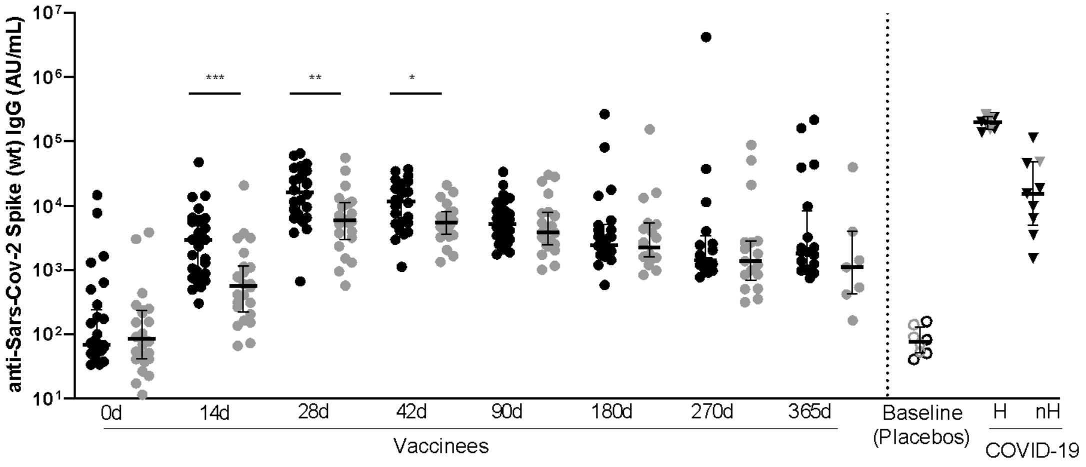
anti-Sars-Cov-2 Spike (wt) IgG (AU/mL)
10^7
10^6
10^5
10^4
10^3
10^2
10^1
***
**
*
0d
14d
28d
42d
90d
180d
270d
365d
Vaccinees
Baseline (Placebos)
H
nH
COVID-19

B

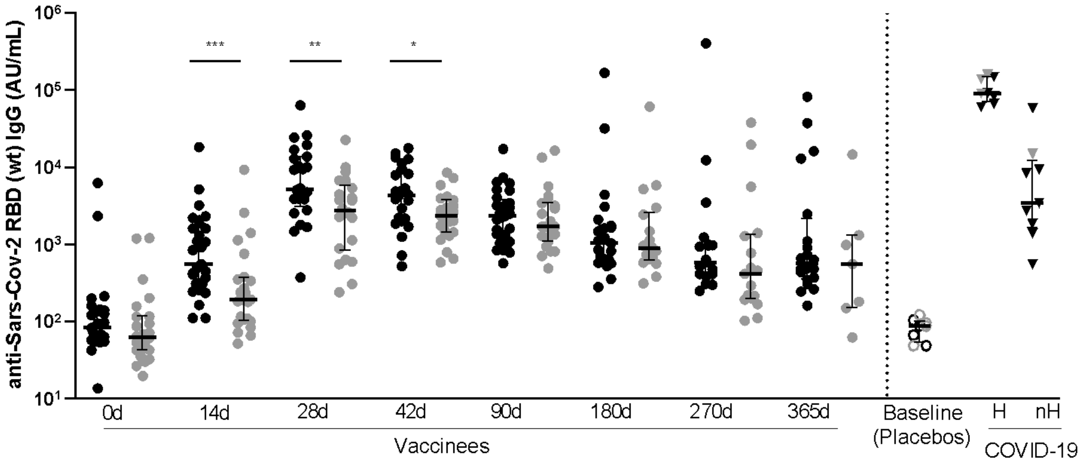
anti-Sars-Cov-2 RBD (wt) IgG (AU/mL)
10^6
10^5
10^4
10^3
10^2
10^1
***
**
*
0d
14d
28d
42d
90d
180d
270d
365d
Vaccinees
Baseline (Placebos)
H
nH
COVID-19

C

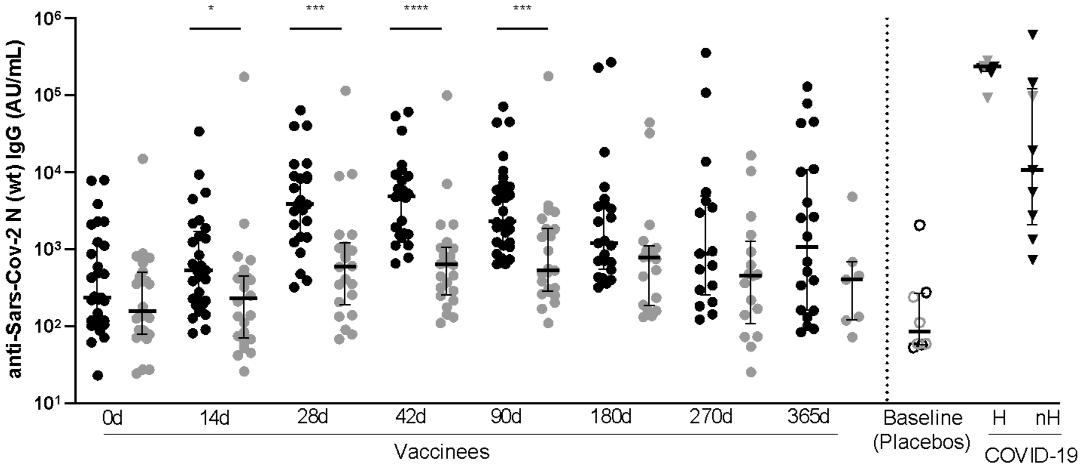
anti-Sars-Cov-2 N (wt) IgG (AU/mL)
10^6
10^5
10^4
10^3
10^2
10^1
*
***
****
***
0d
14d
28d
42d
90d
180d
270d
365d
Vaccinees
Baseline (Placebos)
H
nH
COVID-19

**D**

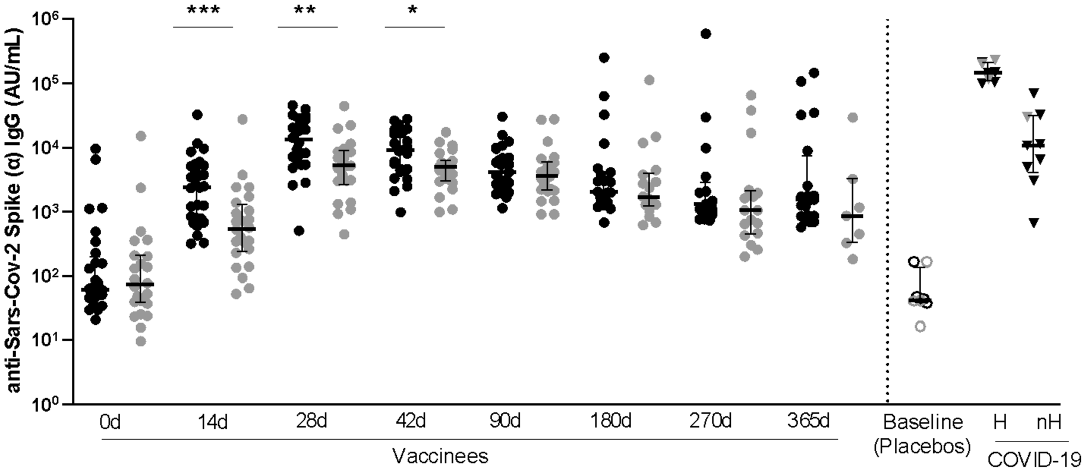
anti-Sars-Cov-2 Spike (α) IgG (AU/mL)
10⁶
10⁵
10⁴
10³
10²
10¹
10⁰
***
**
*
0d
14d
28d
42d
90d
180d
270d
365d
Vaccinees
Baseline
(Placebos)
H
nH
COVID-19

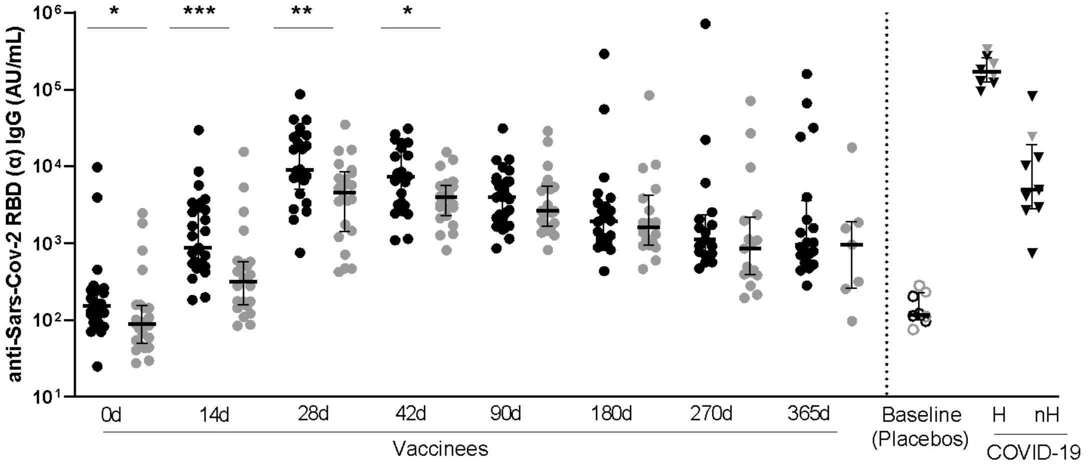
anti-Sars-Cov-2 RBD (α) IgG (AU/mL)
10⁶
10⁵
10⁴
10³
10²
10¹
*
***
**
*
0d
14d
28d
42d
90d
180d
270d
365d
Vaccinees
Baseline
(Placebos)
H
nH
COVID-19

**E**

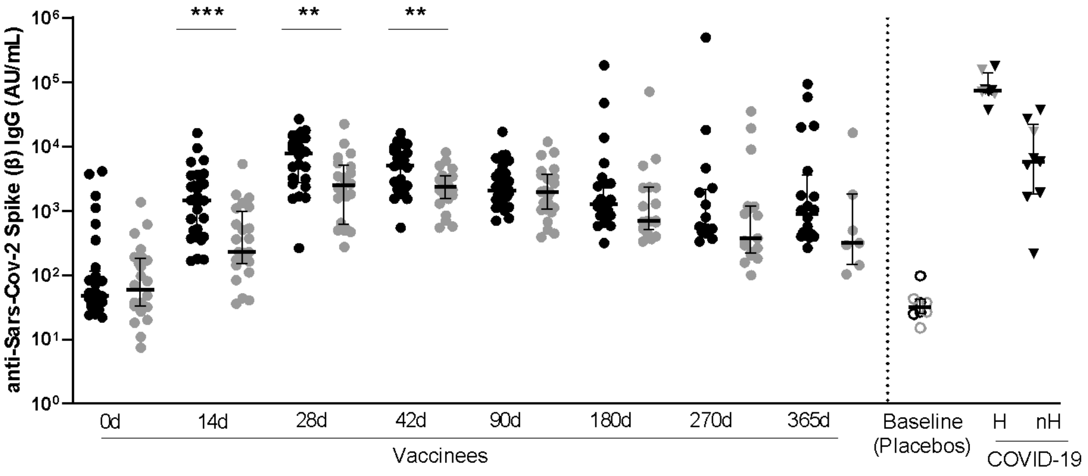
anti-Sars-Cov-2 Spike (β) IgG (AU/mL)
10⁶
10⁵
10⁴
10³
10²
10¹
10⁰
***
**
**
0d
14d
28d
42d
90d
180d
270d
365d
Vaccinees
Baseline
(Placebos)
H
nH
COVID-19

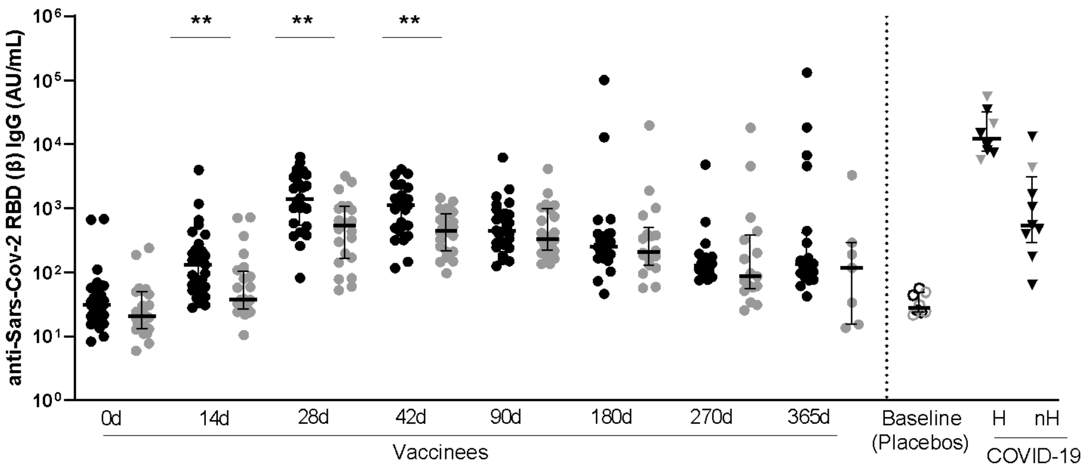
anti-Sars-Cov-2 RBD (β) IgG (AU/mL)
10⁶
10⁵
10⁴
10³
10²
10¹
10⁰
** ** **
0d 14d 28d 42d 90d 180d 270d 365d
Vaccinees
Baseline (Placebos)
H nH
COVID-19

F

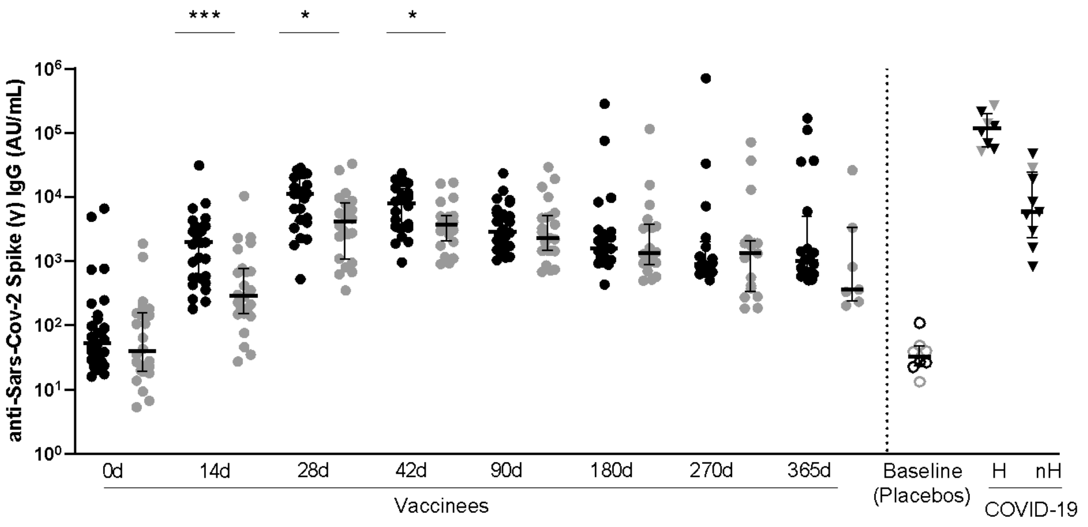
anti-Sars-Cov-2 Spike (γ) IgG (AU/mL)
10⁶
10⁵
10⁴
10³
10²
10¹
10⁰
*** * *
0d 14d 28d 42d 90d 180d 270d 365d
Vaccinees
Baseline (Placebos)
H nH
COVID-19

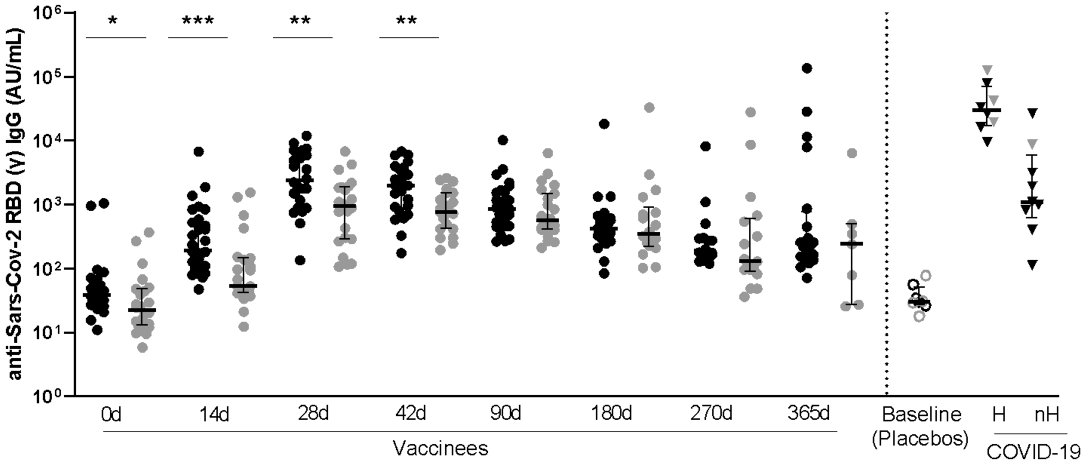
anti-Sars-Cov-2 RBD (γ) IgG (AU/mL)
10⁶
10⁵
10⁴
10³
10²
10¹
10⁰
* *** ** **
0d 14d 28d 42d 90d 180d 270d 365d
Vaccinees
Baseline (Placebos)
H nH
COVID-19

**Figure S1. Specific wt and VOCs SARS-CoV-2 strains IgG titers through one-year follow-up.**

Graphs show anti-SARS-CoV-2 IgG levels in AU/mL from vaccinees throughout one year, and placebos and infected patients as controls. Symbol colours represent age-groups (black: 18-59, grey: ≥60). Symbol shapes represent different volunteer sub-groups (unfilled circle: placebos (n=8), filled circle: vaccinees (n=53), filled triangles: SARS-CoV-2 infected hospitalized 19 days of symptoms onset (median, 17-22 days IQR) (n=6) or non-hospitalized individuals, 46 days of symptoms onset (median, 34-65 days IQR) (n=6)). 0d: vaccinee baseline, day of first vaccine dose. 14d: two-weeks after the first dose, day of the second vaccine dose. Mann-Whitney test was performed to compare both age groups in each visit, and asterisks indicate a statistically significant difference according to the p-value: * $p<0.05$, ** $p<0.01$, *** $p<0.001$, **** $p<0.0001$. IgG levels were measured for (A) wild-type Spike protein; (B) wild-type RBD protein; (C) wild-type nucleocapsid protein; (D) Alpha Spike and RBD proteins; (E) Beta Spike and RBD proteins and (F) Gamma Spike and RBD proteins.

**Table S1.** Demographic data of the study's participants.

| Groups | Age* | Sex | Days between vaccine doses* | Days of symptoms onset* | COVID-19 Diagnosis |
|---|---|---|---|---|---|
| *Placebo (n=8)* | *54 (36-64)* | *62.5% F*<br>*37.5% M* | - | - | - |
| *Vaccinees* | | | | | |
| *18-59 (n=29)* | *36 (31-42)* | *55.2% F*<br>*44.8% M* | *15 (14-18)* | - | - |
| *≥60 (n=24)* | *67 (63-70)* | *33.3% F*<br>*66.7% M* | *16 (14-21)* | - | - |
| *COVID-19 infected individuals* | | | | | |
| *Hospitalized (n=8)* | *53 (43-62)* | *12.5% F*<br>*87.5% M* | - | *19 (17-22)* | *75% PCR*<br>*25% Sorology* |
| *Non-Hospitalized (n=9)* | *40 (33-44)* | *55.6% F*<br>*44.4% M* | - | *46 (34-65)* | *100% PCR* |

**Table S2.** Antibodies and fluorochromes used in the activation induced marker (AIM) assay.

| Fluorochrome | AIM T $CD4^{+}$ cells | AIM T $CD8^{+}$ cells |
|---|---|---|
| BB515 | CD3 (UCHT1) | CD3 (UCHT1) |
| APC-H7 | CD4 (RPA-T4) | CD4 (RPA-T4) |
| PerCP CY5,5 | CD8 (SK1) | CD8 (SK1) |
| BV786 | PD1 (EH12.1) | - |
| APC | CD45RA (HI100) | CD45RA (HI100) |
| PE-CY7 | CD25 (M-A251) | - |
| BV711 | CXCR3 (1C6) | CXCR3(1C6) |
| BV421 | CCR7 (G043H7) | CCR7 (G043H7) |
| Alexa Fluor 700 | CD40/CD154 (24-31) | - |
| APC-R700 | - | CD38 (HIT2) |
| BV510 | CD137 (4B4-1) | CD137 (4B4-1) |
| BV605 | CXCR5 (RF8B2) | - |
| PE | OX40 (Ber-ACT35)* | CD69 (FN50) |
| PE-CF594 | Live-Dead** | Live-Dead** |

*All antibodies were acquired from BD Biosciences, excepting from anti-OX40 (Biolegend). Clones were described in parenthesis. **Cell viability marker, Live/Dead, CF-594, Life Technologies.

**Table S3.** Statistical differences on IgG levels (AU/ml) against SARS-CoV-2 wt and variants of concern (VOCs), Alpha, Beta and Gamma. First line of each protein target represents statistical differences between each age group (18-59 or ≥60) with respective baseline (0d) measures, and the second line represents statistical differences between both age groups (18-59 and ≥60).

| | 0d | | 14d | | 28d | | 42d | | 90d | | 180d | | 270d | | 365d | |
|---|---|---|---|---|---|---|---|---|---|---|---|---|---|---|---|---|
| | 18-59 | ≥60 | 18-59 | ≥60 | 18-59 | ≥60 | 18-59 | ≥60 | 18-59 | ≥60 | 18-59 | ≥60 | 18-59 | ≥60 | 18-59 | ≥60 |
| N | - | - | >0,9999 | >0,9999 | <0,0001 | 0,0756 | <0,0001 | 0,0412 | <0,0001 | 0,0088 | 0,0159 | 0,0383 | 0,1902 | 0,8948 | 0,1004 | >0,9999 |
| | 0,1675 | | 0,0101 | | 0,0003 | | <0,0001 | | 0,0002 | | 0,0562 | | 0,1061 | | 0,1984 | |
| wt Spike | - | - | 0,0004 | 0,2999 | <0,0001 | <0,0001 | <0,0001 | <0,0001 | <0,0001 | <0,0001 | <0,0001 | <0,0001 | 0,0090 | 0,0012 | 0,0010 | 0,1182 |
| | 0,5979 | | 0,0002 | | 0,0047 | | 0,0157 | | 0,4208 | | 0,5239 | | 0,5177 | | 0,2404 | |
| wt RBD | - | - | 0,0015 | 0,3331 | <0,0001 | <0,0001 | <0,0001 | <0,0001 | <0,0001 | <0,0001 | <0,0001 | <0,0001 | 0,0043 | 0,0070 | 0,0008 | 0,1448 |
| | 0,1102 | | 0,0007 | | 0,0095 | | 0,0211 | | 0,5463 | | >0,9999 | | 0,1816 | | 0,4980 | |
| α Spike | - | - | 0,0002 | 0,2779 | <0,0001 | <0,0001 | <0,0001 | <0,0001 | <0,0001 | <0,0001 | <0,0001 | <0,0001 | 0,0068 | 0,0074 | 0,0008 | 0,1919 |
| | 0,7032 | | 0,0004 | | 0,0077 | | 0,0211 | | 0,4533 | | 0,3155 | | 0,1598 | | 0,1455 | |
| α RBD | - | - | 0,0018 | 0,3720 | <0,0001 | <0,0001 | <0,0001 | <0,0001 | <0,0001 | <0,0001 | <0,0001 | <0,0001 | 0,0022 | 0,0037 | 0,0008 | 0,1343 |
| | 0,0188 | | 0,0002 | | 0,0047 | | 0,0188 | | 0,3600 | | 0,8455 | | 0,1705 | | 0,3998 | |
| β Spike | - | - | 0,0003 | 0,1816 | <0,0001 | <0,0001 | <0,0001 | <0,0001 | <0,0001 | <0,0001 | <0,0001 | <0,0001 | 0,0198 | 0,0043 | 0,0019 | 0,2120 |
| | 0,7396 | | 0,0003 | | 0,0067 | | 0,0072 | | 0,2964 | | 0,1615 | | 0,1816 | | 0,1164 | |
| β RBD | - | - | 0,0165 | 0,5523 | <0,0001 | <0,0001 | <0,0001 | <0,0001 | <0,0001 | <0,0001 | <0,0001 | <0,0001 | 0,0253 | 0,0038 | 0,0009 | 0,3004 |
| | 0,0880 | | 0,0023 | | 0,0037 | | 0,0051 | | 0,3897 | | 0,6306 | | 0,2604 | | 0,3698 | |
| γ Spike | - | - | 0,0002 | 0,1953 | <0,0001 | <0,0001 | <0,0001 | <0,0001 | <0,0001 | <0,0001 | <0,0001 | <0,0001 | 0,0083 | 0,0003 | 0,0011 | 0,0854 |
| | 0,3795 | | 0,0002 | | 0,0150 | | 0,0298 | | 0,4315 | | 0,5585 | | 0,7857 | | 0,1036 | |
| γ RBD | - | - | 0,0025 | 0,3257 | <0,0001 | <0,0001 | <0,0001 | <0,0001 | <0,0001 | <0,0001 | <0,0001 | <0,0001 | 0,0205 | 0,0033 | 0,0005 | 0,1286 |
| | 0,0131 | | 0,0002 | | 0,0034 | | 0,0044 | | 0,2558 | | 0,9689 | | 0,2171 | | 0,464 | |

Colours represent statistical difference according to p-value, being grey: no significant, light yellow: * p<0.05, light orange: ** p<0.01, light red: *** p<0.001 and red: **** p<0.0001.
